# Development and Pilot Testing of a Five Item Traumatic Stress Screener for Use with Adolescents in Pediatric Primary Care

**DOI:** 10.1101/2022.02.11.22270757

**Authors:** Lauren C. Ng, Rachel Oblath, Rebecca Brigham, Ming Him Tai, Mandy Coles

## Abstract

**Objectives:** To develop and assess the psychometric properties of the Adolescent Primary Care Traumatic Stress Screen (APCTSS), a five-item yes/no screener for PTSD symptoms in adolescents, and the first developed for pediatric primary care.

**Study Design:** The APCTSS was developed by combining and adapting the UCLA PTSD Reaction Index for DSM-5 with the adult Primary Care PTSD Screen for DSM-5. Adolescent medicine patients were universally approached during clinic visits. With a response rate of 83.6%, 178 patients aged 13-22 (M=18.4, SD=2.3), 64.4% female; 62.1% Black or African-American and 20.7% Hispanic/Latinx, were enrolled. Patients completed APCTSS, Patient Health Questionnaire for Adolescents (PHQ-A), and Child PTSD Symptom Scale for DSM-5 Interview (CPSS-5-I), and 61 completed the Traumatic Events Screening Inventory for Children (TESI-C).

**Results:** 56.7% reported a criterion A trauma, 30.1% met criteria for PTSD, 7.4% met criteria for subsyndromal PTSD, and 19.0% for post-event impairing symptoms. Scores ≥ 2 on the APCTSS optimized sensitivity (.79; 95% CI=.66 to .89) and specificity (.68; 95% CI=.55 to .76) for PTSD, with an area under the curve (AUC) of .79. Sensitivity (.86; 95% CI=.65 to .90), specificity (.77; 95% CI=.60 to .90), and AUC (.86) were even stronger when the TESI-C was used to assess criterion A for PTSD diagnosis. Over half (56.0%) of patients who screened positive on the APCTSS were missed by the PHQ-A.

**Conclusions:** The APCTSS shows promise as an internally consistent, valid, and effective tool for identifying adolescents at high risk of PTSD and traumatic stress.

## Introduction

Almost 80% of adolescents in the US have experienced a traumatic event (Turner et al., 2010), and approximately 7% have post-traumatic stress disorder (PTSD; Merikangas et al., 2010). PTSD is associated with school failure, high-risk sexual behaviors, suicide attempts, substance abuse, relationship problems, involvement in the justice system, and poor physical health outcomes (Greene et al., 2016). PTSD prevalence amongst adult primary care samples ranges from 2% to 39% with a large proportion of heterogeneity explained by differential trauma exposure (Greene et al., 2016). Unfortunately, PTSD and traumatic stress symptoms are routinely unidentified in pediatric primary care (Gerson & Rappaport, 2013), and most pediatricians report that they lack adequate knowledge, skills, and comfort to discuss PTSD, and only 10% assess and treat PTSD (Banh et al., 2008).

Some of the difficulty with identifying patients with traumatic stress symptoms in primary care may stem from the fact that typically individuals with PTSD symptoms do not spontaneously report their mental health symptoms or trauma histories (Read et al., 2006; Wurr & Partridge, 1996). If health providers do not explicitly assess these symptoms, they are usually missed. Indeed, PTSD detection rates in routine adult primary care have found detection rates from 0% to a high of 52% (Greene et al., 2016). Therefore, screening for PTSD may be necessary to detect patients who are coping with trauma-related symptoms. Luckily, research with adult samples has found that most patients are comfortable reporting trauma-exposure and PTSD symptoms on primary care screeners (Goldstein et al., 2017). Recently, adverse childhood experiences (ACES) are being more routinely assessed in pediatric primary care (California Department of Health Care Services, 2021). However, fewer than 20% of trauma exposed youth will develop PTSD (Alisic et al., 2014), thereby reducing the utility of screening for ACES to identify youth most in need of trauma-focused mental health care.

Additionally, there is a lack of validated and feasible assessments for assessing traumatic stress symptoms in pediatric primary care. Primary care requires brief and simple tools (Löwe et al., 2005). However, most PTSD screeners for youth and adults have been developed and validated in specialty mental health with clinical populations and have 17 or more items and multiple response options (Foa et al., 2018; Steinberg et al., 2013). A few brief measures with yes/no response options have been developed, but they predict future PTSD (Kenardy et al., 2006) or assess acute stress, not PTSD (Kassam-Adams & Marsac, 2016). The Child Trauma Screen (CTS) is the only other PTSD screener for children and adolescents that has been validated for use in primary care for adolescents (Lang & Connell, 2018; Lang et al., 2021a; Lang et al., 2021b). However, the CTS was developed in a community mental health clinic and consists of four dichotomous trauma exposure items and six reaction items measured on a 4-point Likert scale (Lang & Connell, 2017). The only PTSD symptom scales developed for, and validated in, primary care have been for adults (Spoont et al., 2013).

Few studies have investigated the implementation and feasibility of PTSD screening in primary care, but studies suggest that even the longest screening tools take only 10 minutes to complete (Spoont et al., 2013). However, given the very limited time available in busy primary care practices, providers and researchers tend to prioritize brevity (Brewin, 2005), and often one or two item screeners, such as the PHQ-2 (Kroenke et al., 2003), have more uptake in routine care. However, one or two item PTSD screeners such as the Single-item PTSD Screener (SIPS; Gore et al., 2008) and the PCL-2 (Lang et al., 2012) have been found to have weak psychometric properties due to limited variation in response options (Spoont et al., 2013). In contrast, the five-item Primary Care PTSD Screen (PC-PTSD) has reasonable psychometric properties (Prins et al., 2016), and is widely used and accepted (Spoont et al., 2015). The characteristics of the PC-PTSD that are attractive for use in primary care include: (1) being self-report, (2) being only 4 or 5 items long, (3) focusing on meaningful empirically supported symptoms, (4) not utilizing Likert-style responses, (5) not requiring an interview about trauma exposure, (6) focusing on current PTSD, and (7) having psychometrics from primary care (Prins et al., 2003).

Given that a wide range of stress-related symptoms in childhood confer a transdiagnostic diathesis for mental disorders including, but not limited to, PTSD (Basu et al., 2020; D’Andrea et al., 2012), somatic syndromes (Afari et al., 2014), and poor physical health (O’Donovan et al., 2011) in later adolescence and adulthood, we sought to develop a scale that would not only identify adolescents who meet DSM-5 PTSD diagnostic criteria (American Psychiatric Association, 2013), but also those with any functionally-impairing post-trauma symptoms, including subsyndromal PTSD (Alessi et al., 2013). Individuals with subsyndromal PTSD experience substantial functional difficulties that require mental health treatment (Pietrzak et al., 2012) and often progress to full PTSD (Fink et al., 2018). The need to identify youth with post-trauma symptoms who might not meet full diagnostic criteria for PTSD may be particularly pertinent when trying to identify high-risk youth who may be expected to present with less severe symptoms than individuals detected through routine clinical care (Sonis, 2013). Additionally, we sought to identify patients with trauma-related symptoms due to a non-DSM-5 Criterion A trauma such as relationship difficulties, non-violent deaths of loved ones, bullying, and separation from parents. PTSD symptoms following non-Criterion A trauma are often similar to, or sometimes worse than, those reported after Criterion A traumas (Lansing et al., 2017). Early identification and treatment of all of these adolescent patients may prevent more severe problems and is therefore of interest to pediatrics providers.

The three aims of this study were to (1) develop the Adolescent Primary Care Traumatic Stress Screen (APCTSS), a screener that is feasible and acceptable for pediatrics, (2) pilot the APCTSS in an adolescent medicine clinic, and (3) assess its psychometric properties, including the internal structure of the scale, its concurrent and discriminant validity, and its sensitivity/specificity and clinical cut point for identifying PTSD, subsyndromal PTSD, or clinically impairing symptoms associated with a non-Criterion A distressing event.

## Method: Aim 1 - Development of the APCTSS

### Study Site

This study was conducted in [edited out for blind review], which treats patients aged 12-22 years old. [edited out for blind review] is the largest safety-net hospital in [edited out for blind review], with 57% percent of patients from underserved populations and 72% insured by government payors ([edited out for blind review]). This study was approved and overseen by the [edited out for blind review] IRB (#).

### Procedures

The 31-item UCLA PTSD Reaction Index for DSM-5 (UCLA-RI-5; Steinberg et al., 2013), a well-validated PTSD scale for children and adolescents, was adapted and combined with the 5-item adult Primary Care PTSD Screen for DSM-5 (PC-PTSD-5; Prins et al., 2016). Six attending-level healthcare providers in the BMC Departments of Psychiatry (two psychologists and one psychiatrist) and Pediatrics (one adolescent medicine physician and two social workers) with expertise in diagnosing and treating PTSD in adolescents independently identified UCLA-RI-5 items corresponding to each of the items on the PC-PTSD-5.

The results of the mapping exercise were presented to five clinicians in the BMC Department of Psychiatry who are experts in diagnosing and treating PTSD in adolescents (three psychiatrists and two psychologists), of which three also participated in the item mapping. The five experts reached consensus on which of the identified UCLA-RI-5 items they thought best represented the symptom profile of PTSD in adolescents and best discriminated adolescents with PTSD from those with other mental health disorders (See Table 1 for consensus results). The first and fourth authors refined and adapted the item language and instructions to develop an introductory prompt.

**Table 1.**
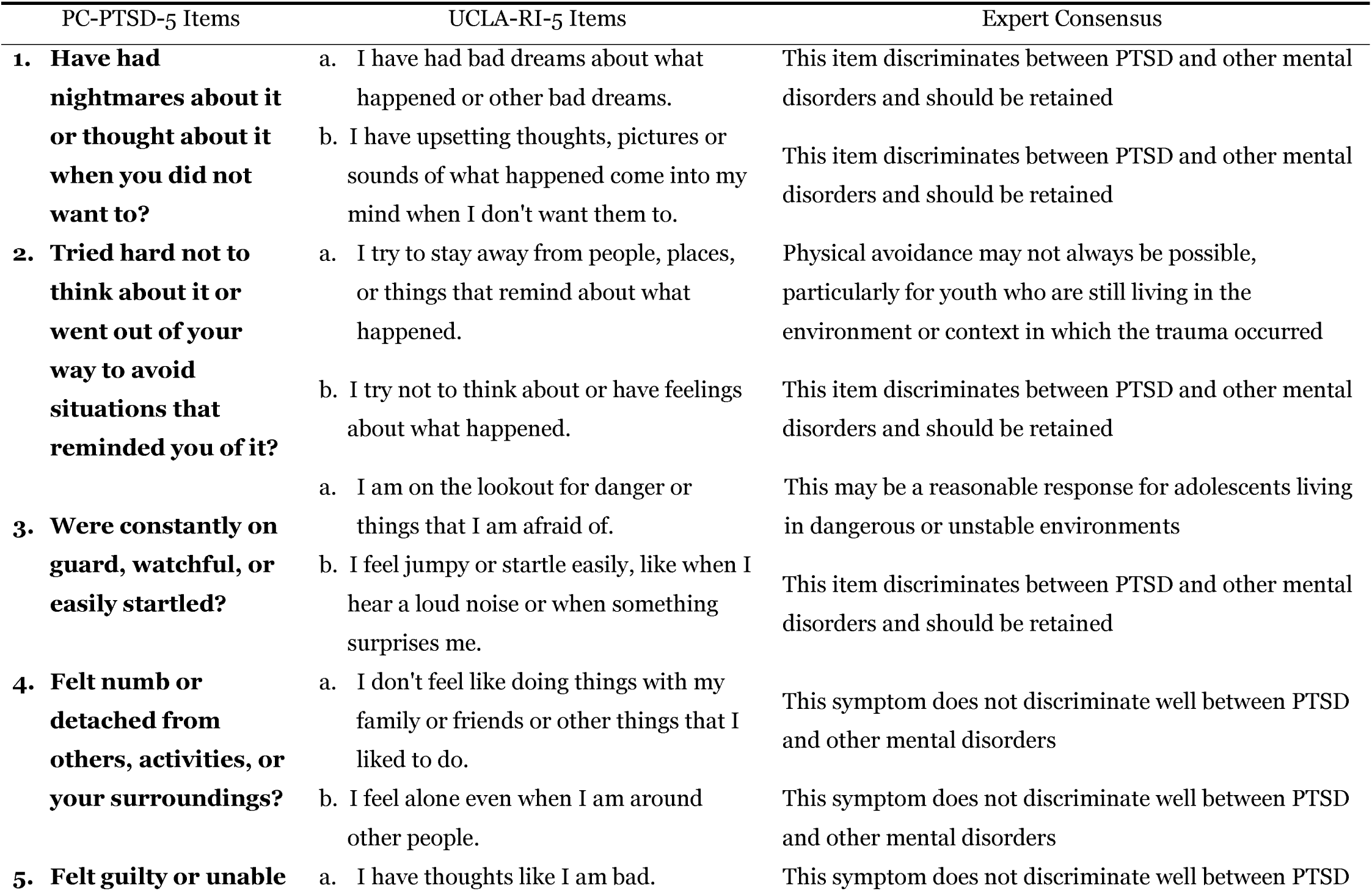
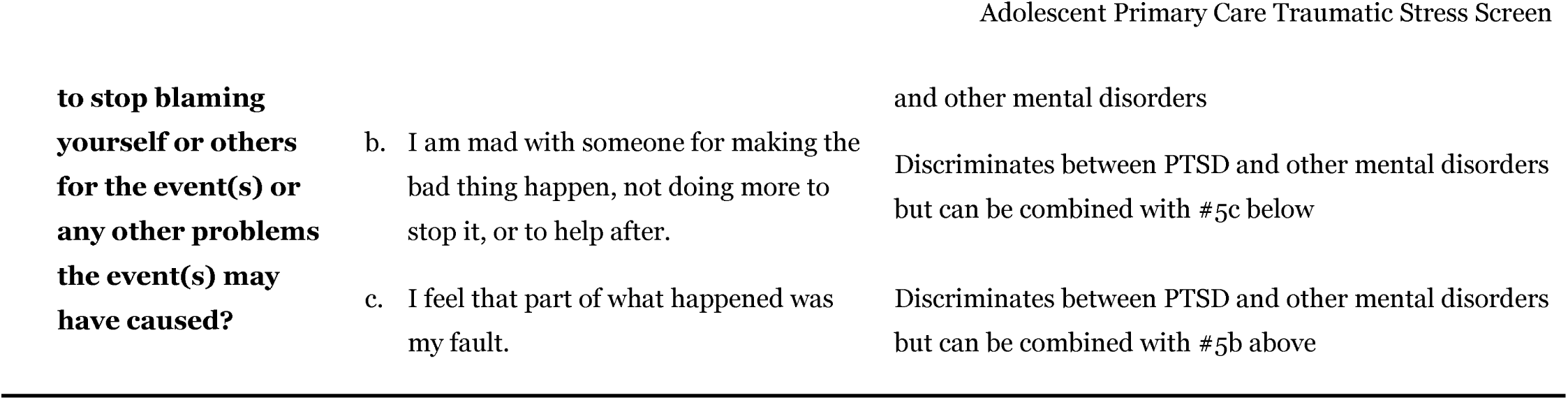
Items from the UCLA-RI-5 that corresponded to the PC-PTSD-5, as rated by expert clinicians PC-PTSD-5 Items UCLA-RI-5 Items Expert Consensus.

| PC-PTSD-5 Items | UCLA-RI-5 Items | Expert Consensus |
| --- | --- | --- |
| **1. Have had nightmares about it or thought about it when you did not want to?** | a. I have had bad dreams about what happened or other bad dreams. | This item discriminates between PTSD and other mental disorders and should be retained |
|  | b. I have upsetting thoughts, pictures or sounds of what happened come into my mind when I don't want them to. | This item discriminates between PTSD and other mental disorders and should be retained |
| **2. Tried hard not to think about it or went out of your way to avoid situations that reminded you of it?** | a. I try to stay away from people, places, or things that remind about what happened. | Physical avoidance may not always be possible, particularly for youth who are still living in the environment or context in which the trauma occurred |
|  | b. I try not to think about or have feelings about what happened. | This item discriminates between PTSD and other mental disorders and should be retained |
| **3. Were constantly on guard, watchful, or easily startled?** | a. I am on the lookout for danger or things that I am afraid of. | This may be a reasonable response for adolescents living in dangerous or unstable environments |
|  | b. I feel jumpy or startle easily, like when I hear a loud noise or when something surprises me. | This item discriminates between PTSD and other mental disorders and should be retained |
| **4. Felt numb or detached from others, activities, or your surroundings?** | a. I don't feel like doing things with my family or friends or other things that I liked to do. | This symptom does not discriminate well between PTSD and other mental disorders |
|  | b. I feel alone even when I am around other people. | This symptom does not discriminate well between PTSD and other mental disorders |
| **5. Felt guilty or unable** | a. I have thoughts like I am bad. | This symptom does not discriminate well between PTSD |

## Results: Aim 1 - Development of the APCTSS

At least four of the six raters believed that 11 items from the UCLA-RI-5 corresponded to at least one of the five items on the PC-PTSD-5 (See Table 2). There was consensus that both of the UCLA-RI-5 items that corresponded to PC-PTSD-5 item #1 (nightmares) were more likely to be present in adolescents with PTSD than adolescents with other mental disorders, and these items were retained. For PC-PTSD-5 item #2 (avoidance), the UCLA-RI-5 item describing imaginal rather than physical avoidance was selected, since many adolescents are unable to physically avoid reminders of trauma. To represent PC-PTSD-5 item #3 (hyperarousal) they selected an item that did not include being on the lookout for danger, as this may be a common response for non-symptomatic adolescents exposed to potentially dangerous environments. The experts agreed that the two UCLA-RI-5 items that corresponded to PC-PTSD-5 item #4 (numbing/detachment) did not adequately differentiate PTSD from other mental disorders in adolescents, and neither were retained. Two of the UCLA-RI-5 items that corresponded to PC-PTSD-5 item #5 (guilt/blame) were deemed to discriminate between PTSD and other mental disorders and were combined into one. The UCLA-RI-5 item “I have thoughts like I am bad” was not retained due to strong overlap with symptoms of depression and anxiety. The initial prompt for the APCTSS was adapted from the UCLA-RI-5 self-report trauma history screener (See Figure 1 for the complete APCTSS measure).

**Table 2.** Item-level descriptive statistics for the APCTSS.

| Item | N | Percentage of<br>participants endorsing<br>item |
| --- | --- | --- |
| Bad dreams about scary experiences or other bad dreams | 74 | 41.60% |
| Upsetting thoughts, pictures or sounds of scary experiences come into your mind when you didn't want them to | 61 | 34.30% |
| Tried not to think about or have feelings about scary experiences | 70 | 39.30% |
| Mad at yourself or someone else for making the scary experiences happen, not doing more to stop it, or to help after | 33 | 18.50% |
| Felt jumpy or easily startled, like when you hear a loud noise or when something surprises you? | 79 | 44.40% |

**Figure 1.**
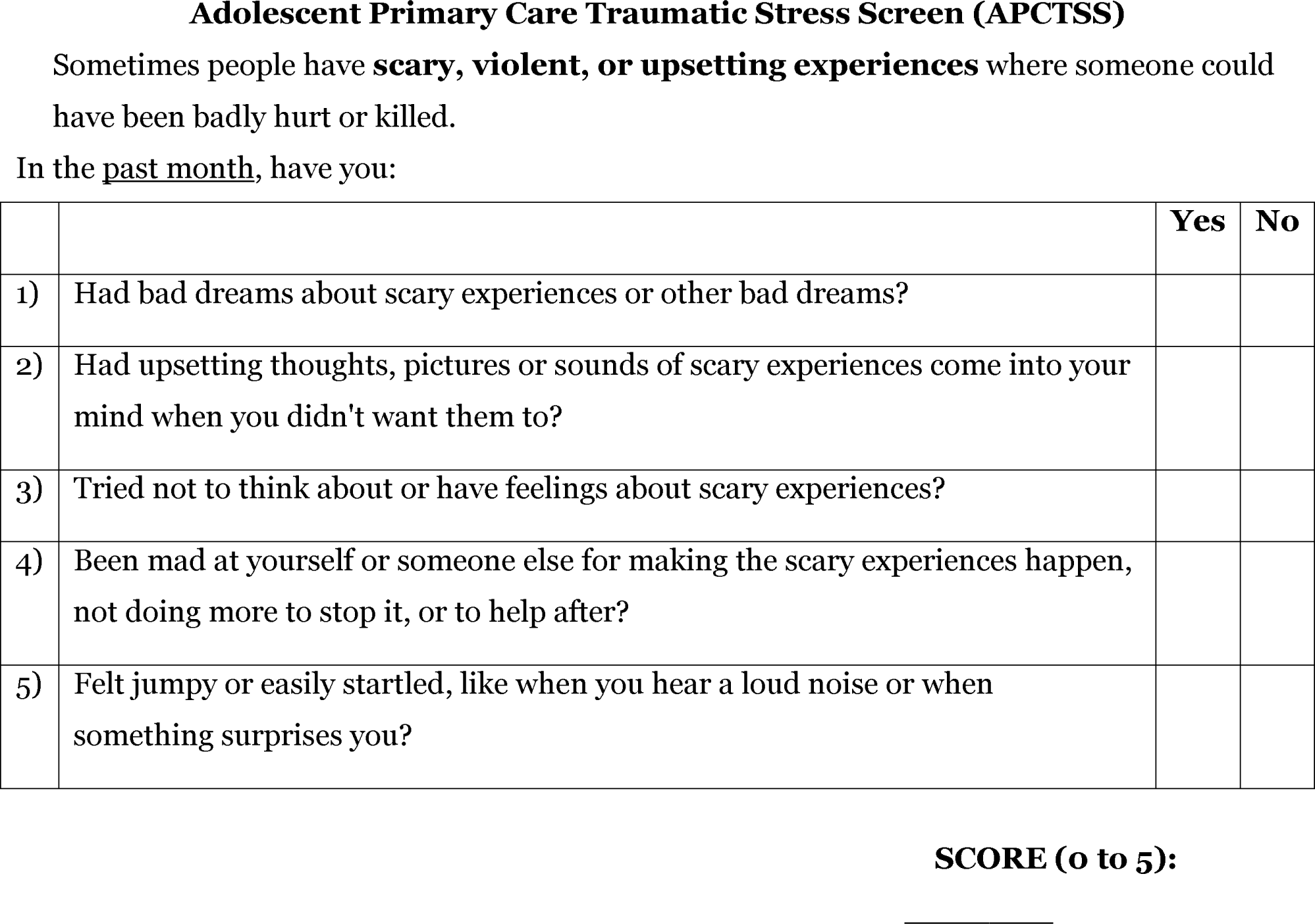
The Adolescent Primary Care Traumatic Stress Screen (APCTSS)

### Method: Aim 2 - Assessment of the Psychometric Properties of the APCTSS Recruitment

When a researcher was available for assessment, all adolescent medicine patients seen in the clinic were informed about the study and invited to participate. Patients over 18 provided verbal informed consent, while patients under 18 and a legal guardian provided verbal assent and consent, respectively. Recruitment took place in the waiting room and in examination rooms while patients were waiting to be seen by clinic staff. Informed consent and completion of study measures took place either in a private therapy office or in an examination room.

### Participants

Participants were 178 patients aged 13-22 (M = 18.4, SD = 2.3) who visited the clinic between December 2018 and February 2020. The majority (83.6%) of patients approached agreed to participate, and 4.7% were excluded due to being unable to complete the study requirements in English. Almost two-thirds of the sample were female (64.4%), 23.6% were male, and 12.0% were transgender or non-binary. In terms of race/ethnicity, 62.1% of participants identified as Black or African-American, 20.7% identified as Hispanic or Latino, 9.4% identified as White or Caucasian, and 7.4% identified as another race/ethnicity or as multi-racial. Sample demographics were reflective of the overall clinic population.

### Sample Size and Power

The sample size goal was 200 participants based on a power calculation assuming a sample prevalence rate of PTSD of 30%, a sensitivity of 0.85 and specificity of 0.90 with 95% CI half widths of 0.09 for sensitivity and 0.07 for specificity. We had collected data on 178 of 200 participants by the end of February 2020, when data collection was stopped due to Covid-19. Due to uncertain timeline of resumed in-person data collection, we concluded data collection at 178 participants.

### Measures

The APCTSS was developed in earlier phases of the study and described above. Self-reported depression was assessed using the Patient Health Questionnaire for Adolescents (PHQ-A;, Johnson et al., 2002), a nine-item survey designed for use in primary care with adolescents aged 13-18. The Traumatic Events Screening Inventory for Children (TESI-C; Ford et al., 2002) assessed potentially traumatic events. Participant responses were classified as Criterion A traumatic events in accordance with DSM-5 guidelines (American Psychiatric Association, 2013; Pai et al., 2017). PTSD symptoms were assessed using the Child PTSD Symptom Scale for DSM-5 Interview (CPSS-5-I; Foa et al., 2018). which has been validated with youth between the ages of 8 and 18. Cronbach’s alpha of the symptom scale was .95 in this sample. The CPSS-5-I interview begins with a prompt asking participants to “tell me about the most upsetting or scary experience you’ve ever had” and then provides examples (Foa et al., 2018). Interviewers recorded the participant’s most upsetting experiences and when they occurred, and also separately recorded whether the experiences included actual or threatened death, serious injury, or sexual violation. After recording the most upsetting event, the interviewers used the CPSS-5-I to assess the frequency of 20 PTSD symptoms that participants experienced in the past month related to the most upsetting event. Symptoms were rated on a 5-point Likert scale (0=not at all; 4=6 or more times a week/almost always). Finally, the CPSS-5-I assesses the frequency of functional impairment due to PTSD symptoms across seven domains using the same Likert scale.

Participants were classified as having PTSD if they reported a Criterion A trauma and at least all of the following: one intrusion item, one avoidance item, two changes in cognition and mood items, two increased arousal and reactivity items, and three impairment items (American Psychiatric Association, 2013; Foa et al., 2018). Participants were classified as having sub-syndromal PTSD if they endorsed a Criterion A trauma and at least two symptom categories and one functional impairment item (American Psychiatric Association, 2013). Participants were classified as having post-distressing event symptoms if they endorsed at least two symptom categories and at least one functional impairment item, but did not report a qualifying Criterion A trauma.

### Study Procedures

After informed consent, participants independently completed the APCTSS and PHQ-A with pencil and paper, which is the typical form of PTSD screener administration (Brewin, 2005; Spoont et al., 2013) and which is also the standard screening approach in the clinic. Upon completion of the two screeners, two researchers members, who were a PhD candidate in Applied Human Development and a clinical psychology master’s student, verbally administered the CPSS-5-I as an interview (Foa et al., 2018) and participants completed a demographic questionnaire on their own using pencil and paper. The researchers were blind to the participant responses on the APCTSS and the PHQ-A.

During data collection, researchers suspected that some participants who reported non-Criterion A events as their most upsetting experience in response to the open-ended prompt may have also experienced a Criterion A trauma. Therefore, after the first 9 months of data collection, we started administering the TESI-C (Ford et al., 2002), a trauma events checklist, to evaluate whether participants who completed an inventory of traumatic events reported the same rates of Criterion A trauma as those who were just asked to share the most scary or upsetting event they had experienced when prompted during the CPSS-5-I. The initial open-ended prompt on the CPSS-5-I about “the most upsetting or scary experiences you’ve ever had” was asked first, then researchers verbally administered the TESI-C, and then the researchers administered the remaining PTSD symptoms and functional impairment items of the CPSS-5-I. Approximately one-third of participants (N = 61) had the TESI-C verbally administered after the CPSS-5-I open-ended prompt, and prior to the CPSS-5-I symptom and functional impairment assessment. All participants received a $5 cash card for their participation.

### Statistical analysis

Analysis included examining (1) the internal structure of the APCTSS using Cronbach’s alpha, (2) concurrent and discriminant validity using Pearson’s correlation coefficients with the CPSS-5-I and the PHQ-A, and (3) sensitivity and specificity to differentiate participants with and without PTSD and optimal diagnostic cutoff scores for the APCTSS through the use of receiver operating characteristic (ROC) analysis and calculation of the Youden index. In addition, we compared the ability of the APCTSS to identify adolescents at risk of subsyndromal PTSD symptoms or undetected trauma exposure who would otherwise not be identified by the PHQ-A. Post-hoc ROC analyses were run on the subset of participants who completed the TESI-C to assess area under the curve (AUC), sensitivity, and specificity when accounting for trauma disclosed on the TESI-C when assessing Criterion A for PTSD diagnosis.

### Results: Aim 2 - Assessment of the Psychometric Properties of the APCTSS Descriptive Statistics

Over half of the sample (57.3%) endorsed at least one item on the APCTSS, and 39.3% endorsed two or more items (See Table 2 for item-level descriptives). More than half (56.7%) of participants reported experiencing a criterion A trauma on either the TESI-C or CPSS-5-I. Participants who completed the TESI-C were more likely to report Criterion A trauma experiences (71.2%) than those who did not (49.6%), χ2(1) = 7.50, p < .01. The average score on the CPSS-5-I symptom scale was 20.28 (SD = 17.60) regardless of whether they had a qualifying criterion A trauma, indicating that the average BMC adolescent medicine primary care patient endorsed moderate levels of PTSD symptoms. Almost one-third (30.1%) met criteria for PTSD, 7.4% met criteria for subsyndromal PTSD, and an additional 19.0% met criteria for post-event impairing symptoms. The average score on the PHQ-A was 5.80 (SD = 5.39), indicating non-clinical levels of depression symptoms. Results of Fisher’s exact tests indicated that there were no differences in PTSD rates (Fisher’s exact = 0.78), at least subsyndromal PTSD (Fisher’s exact = 1.67), or at least impairing symptoms (Fisher’s exact = 1.23) by race/ethnicity, although we were underpowered to detect differences for White or multiracial or other race youth.

### Internal Consistency

The Cronbach’s alpha coefficient for the APCTSS was .77, which is considered adequate. Coefficient alpha did not improve significantly with the removal of any of the 5 items.

### Sensitivity, Specificity, and Positive and Negative Predictive Value

ROC analysis and the Youden index score indicated that a score of 2 or higher on the APCTSS was associated with optimal values for sensitivity (.79; 95% CI=.66 to .89) and specificity (.68; 95% CI = .59 to .76) for PTSD diagnosis (see Table 3). A cut-off score of 2 yielded a positive predictive value (PPV) of .55 (95% CI = .47 to .62) and a negative predictive value (NPV) of .87 (95% CI = .80 to .92). The area under the curve (AUC) was .79. A cut-off score of 2 was also the optimal cut-off for detecting sub-syndromal PTSD, with a sensitivity of .78 (95% CI=.67 to .88), specificity of .73 (95% CI=.64 to .82), AUC of .81, and a Youden index of .51. A cut-off score of 2 resulted in 16 false positives (9.0%; i.e., youth identified as being at high risk on the screener, but did not have PTSD, sub-syndromal PTSD, or post-event impairing symptoms), but missed 32 youth (18.0%) who had impairing symptoms. Comparatively, a score of 1 resulted in 41 (23.0%) false positives but only missed 12 (6.7%) youth who did have these symptoms, of which four had PTSD.

**Table 3.** Number and Percent of participants with PTSD diagnoses and symptoms compared to APCTSS scores.

| APCTSS Score | 0 | 1 | 2 | 3 | 4 | 5 |
| --- | --- | --- | --- | --- | --- | --- |
|  | n (%) | n (%) | n (%) | n (%) | n (%) | n (%) |
| Participants with score | 53 (29.8%) | 45 (25.3%) | 24 (13.5%) | 20 (11.2%) | 16 (9.0%) | 20 (11.2%) |
| Participants with score AND PTSD | 4 (7.6%) | 7 (15.6%) | 9 (37.5%) | 10 (50.0%) | 7 (43.8%) | 16 (80.0%) |
| Participants with score AND probable subsyndromal PTSD | 0 (0%) | 3 (6.67%) | 3 (12.5%) | 2 (10%) | 2 (12.5%) | 2 (10%) |
| Participants with score AND post-event impairing symptoms (non-Criterion A event) | 8 (15.1%) | 10 (22.2%) | 4 (16.7%) | 3 (15.0%) | 5 (31.3%) | 1 (5%) |
| Participants with the score and either PTSD, subsyndromal PTSD, or post-event impairing symptoms (non-Criterion A event) | 12 (22.6%) | 20 (44.4%) | 16 (66.7%) | 15 (75.0%) | 14 (87.5%) | 19 (95.0%) |
| Participants without post-event or post-trauma impairing symptoms | 41 (77.3%) | 25 (55.6%) | 8 (33.3%) | 5 (25.0%) | 2 (12.5%) | 1 (5.0%) |

Results of the post-hoc ROC analyses were run on the subset of participants who completed the TESI-C to assess AUC, sensitivity, and specificity when accounting for Criterion A trauma exposure that was not reported in response to the open-ended prompt on the CPSS-5-I. A cut-off score of 2 was also the optimal cut-point for the subset of participants who completed the TESI-C, but sensitivity (.86; 95% CI=.65 to .90), specificity (.77; 95% CI=.60 to .90), PPV (.70; 95% CI = .56 to .82), NPV (.90; 95% CI = .76 to .96) and AUC (.86) were stronger for PTSD diagnosis compared to the full sample. Similar results were observed for subsyndromal PTSD and post-event impairing symptoms (See Table 3 for complete results).

### Concurrent Validity

The APCTSS was strongly correlated with the total CPSS-5-I symptom score (r = .71, p < .001) and the total score for impairment items (r = .62, p < .001).

### Discriminant Validity

The APCTSS was moderately correlated with the PHQ-A (r = .55, p <. 001), and the association was significantly lower than the association between the APCTSS and the CPSS-5-I (z = 3.87, p < .001; Steiger, 1980). Over half of patients (56.0%) who screened positive on the APCTSS (score ≥ 2) would not have been flagged using the PHQ-A, including 60.8% of patients who had probable PTSD, subsyndromal PTSD, or post-event impairing symptoms.

## Discussion

The need for a feasible and valid primary care PTSD screener for pediatrics is high. In this sample, more than 55% of adolescents presenting for primary care reported experiencing a criterion A trauma, 30% met DSM-5 diagnostic criteria for PTSD, and a further 26% either had sub-syndromal PTSD or post-event impairing symptoms. All together, 56% had functionally impairing symptoms associated with a traumatic or difficult event. Although the PTSD prevalence rate found in this study was high, results also suggest that our study may have actually underestimated the prevalence of PTSD in our sample. Because more participants who were administered the TESI-C endorsed criterion A traumas than participants who were only asked to report trauma in response to an open ended prompt, it is possible that the false positive rate on the screener was artificially inflated because some of those false positives would have qualified for PTSD or sub-syndromal PTSD had we administered the TESI-C, since more of them likely would have endorsed criterion A experiences. The finding of somewhat higher trauma reports using a trauma checklist has been observed in other studies that specifically sought to answer this question (Monson et al., 2016). Therefore, it is likely that more than 30% of the adolescent patients seen in routine pediatric primary care at BMC have PTSD, which is often not detected. The high rates of PTSD and post-trauma symptoms identified by this study emphasize the need to have a psychometrically sound and feasible traumatic stress screener for use in pediatric primary care, particularly clinics serving a low-income and BIPOC youth.

The PTSD prevalence in this sample is much higher than rates found from epidemiological studies (Merikangas et al., 2010), but is similar to other populations of youth who receive primary care services in safety net hospitals or federally qualified health centers (Selwyn et al., 2019) which serve a higher proportion of patients who are racial/ethnic minorities, non-English speaking, uninsured, underinsured, undocumented, or low-income (Gaskin & Hadley, 1999; Lasser et al., 2021; Nath et al., 2016). Our study sample was more than 60% Black or African-American and more than 21% Hispanic or Latino, which mirrors the overall BMC pediatric primary care clinic population. Although we did not collect data on income or socioeconomic status, more than 70% of BMC patients are insured by government payors (Boston Medical Center, 2016), a blunt proxy for low-income status. Low-income and BIPOC (black, indigenous, and people of color) youth are disproportionately affected by traumatic and adverse events (Giano et al., 2020; Mersky et al., 2021), which are at least partially downstream outcomes of structural inequities. Indeed, structural inequities arise from exploitative power dynamics (Laster Pirtle, 2020; Prins et al., 2021), as do most types of trauma. Prejudice, discrimination, and systemic or structural inequities such as policies, practices, and behaviors that perpetuate disparities in wealth, housing, education, employment, healthcare, and opportunities can be traumatic themselves (Mikhail et al., 2018). Moreover, they create conditions that increase the likelihood that low-income and BIPOC youth will be exposed to other traumas, such as violence and death (Knopov et al., 2019; Wilkins et al., 2019). One consequence of the disproportionate exposure to trauma and ACEs that low-income and BIPOC youth cope with is a higher rate of subsequent PTSD and associated mental health symptoms and disorders (Lopez et al., 2017; Price et al., 2019; Pulsifer et al., 2019).

This study developed and pilot tested the Adolescent Primary Care Traumatic Stress Screen (APCTSS), the first PTSD and traumatic stress screener for adolescents developed specifically for pediatric primary care. The development process leveraged expert and stakeholder knowledge by asking pediatricians, psychiatrists, psychologists, and social workers who all provide care to trauma-affected adolescent patients in BMC to identify common post-trauma symptoms expressed by their patients, to refine the language in the measure to be developmentally appropriate, to ensure that items accurately reflect symptoms rather than environmental or social stressors which may disproportionately impact some youth or communities (i.e., not including ‘being on the lookout for danger’”), and to create a feasible and useable measure that would have a very high likelihood of being adopted into routine practice in an urban safety net pediatric primary care clinic. Validity and reliability testing indicated that the APCTSS is internally consistent, has good concurrent and discriminant validity, and is effective at identifying adolescents at high risk for post-trauma symptoms and PTSD.

Our results suggest that a cut-off score of 2 on the APCTSS is appropriate for correctly identifying adolescents who are at high risk of having PTSD or related symptoms and require further assessment. The AUC of .79 was comparable to the AUC rate of other PTSD screeners used in primary care, which range from .75 to .93 (Spoont et al., 2013). and an AUC of 0.7 and higher is generally considered strong in the field of applied psychology (Rice & Harris, 2005). Moreover, a cut-off score of 2 yielded a PPV of .55 (95% CI = .47 to .62) and a NPV of .87 (95% CI = .80 to .92), in line with the performance of the PC-PTSD in in routine care primary care samples (0.41 and 0.97, respectively; Ouimette et al., 2008) and in rigorous validity studies (0.51 and 0.99. respectively; Prins et al., 2016).

While the results of the ROC curve analysis were strong for the full sample, they were even better in the sub-sample of participants who completed the TESI-C, with an AUC of .86, a sensitivity of .86 (95% CI=.65 to .90) and specificity of .77 (95% CI=.60 to .90). The results suggest that the lower scores in the full sample may be better explained by misclassification of some participants with PTSD as not having PTSD rather than inaccurate detection of the APCTSS. It is likely that the true classification accuracy of the APCTSS is closer to an AUC of .86 than .79. Regardless, the APCTSS was quite successful in accurately detecting high risk participants in this sample, demonstrating internal reliability, convergent validity, sensitivity, and specificity comparable to the Child Trauma Screen (CTS), a 10-item measure that uses a four-point Likert scale, and has also been validated (but not developed) in pediatric primary care (Lang et al., 2021b). However, the brevity, simplicity, and lack of a traumatic events scale of the APCTSS, along with its similar psychometric properties to longer screeners evaluated in pediatric primary care, may increase its likelihood of being adopted as the measure of choice in routine pediatric primary care, just as the PC-PTSD-5 has been adopted in routine adult primary care (Bovin et al., 2021).

Notably, almost 70% of patients who screened positive on the APCTSS would not have been detected by the PHQ-A (Johnson et al., 2002). This included over half who had PTSD, subsyndromal PTSD, or post-event impairing symptoms. Reliance solely on generalized distress and depression scales to screen for common mental disorders may miss more than 50% of adolescents with trauma-related distress and impairment, similar to findings with adult samples (Ouimette et al., 2008). Including a brief screener for PTSD symptoms may identify youth coping with PTSD who would otherwise not receive care.

Researchers have noted that the evidence base for universal screening of PTSD in primary care is limited, and therefore universal screening is not yet warranted. However, they suggest that targeted screening of patients who spontaneously report PTSD symptoms (Sonis, 2013) or trauma exposure, have other mental health or substance abuse problems, or who are non-responsive to treatment for insomnia or pain (Greene et al., 2016), should be screened for trauma-related symptoms. This approach may be more cost-effective and would be expected to result in higher specificity and sensitivity (Sonis, 2013). The APCTSS could help fill the gap in pediatric primary care by providing a tool to quickly screen these high-risk patients.

Results must be interpreted within the limitations of the methods, including the reliance on youth self-report. However, the goal was to identify adolescents using self-report methods in assessing the APCTSS, and adolescents tend to be accurate reporters of internalizing distress (Weems et al., 2005), and so we concluded that self-report assessments would be appropriate. A second limitation was the lack of a clinician administered structured clinical interview, although we did utilize the interview version of the CPSS-5. Studies have found correlations exceeding .90 between self-report PTSD symptom scales and clinician rated structured interviews (Blanchard et al., 1996), and so we primarily utilized self-report measures for ease of administration. Future studies should validate the APCTSS using a clinician-rated structured clinical interview.

This study is also limited by the lack of inclusion a trauma event checklist for most of the participants. The inclusion of the TESI-C for the last 34% of participants allowed us to identify more participants that qualified for PTSD or subsyndromal PTSD, and it is likely that we misclassified earlier participants as having post-event impairing symptoms when they met criteria for PTSD or subsyndromal PTSD. A further limitation is that our sample size did not allow us to look at psychometric properties by race/ethnicity or gender, and we did not include a measure of socioeconomic status. Future studies should utilize a larger sample size to assess differential validity or reliability across different demographic variables.

In addition, there was overlap of expert participants during the two phases of the development of the measure, such that three experts participated in step one, which was independently identifying UCLA-RI-5 items that corresponded to PC-PTSD-5 items, and also participated in step two, which was the consensus discussion of the results from step one to select specific items for inclusion on the APCTSS. It may be that the overlap in participants decreased the variance in expert opinion or biased the results in step two. However, we believe that use of both an independent rating and a consensus rating also strengthened item selection.

Finally, the APCTSS was developed and validated in the BMC Department of Pediatrics’ adolescent medicine clinic and results may not be generalizable to other clinics or populations. However, the purpose was to develop a measure that was contextually and culturally appropriate for a busy urban safety net pediatrics clinic and its patients. We believe that the resulting measure, which capitalizes on expert and stakeholder feedback, UCLA-RI-5 and PC-PTSD-5 items, and reliability and validity assessed in routine care in a diverse population makes it potentially generalizable to many clinics. The study also had several other strengths which have typically been limitations of the other studies of primary care PTSD screeners (Spoont et al., 2013), including universally approaching and enrolling primary care patients, non-selective recruitment for CPSS-5-I administration, and conducting the CPSS-5-I without knowledge of the results of the APCTSS.

## Conclusion

Many youth with trauma-related mental health symptoms and functional impairment have not been identified in pediatric primary care. This missed opportunity for early identification, prevention, and intervention, may have contributed to a host of poor outcomes for these youth. The development of an effective and feasible PTSD screening tool for youth primary care may improve the health and well-being for some of our most vulnerable adolescents.

## Data Availability

The deidentified datasets generated during and analyzed during the current study are available from the corresponding author upon reasonable request.

